# Derivation, External Validation and Clinical Implications of a deep learning approach for intracranial pressure estimation using non-cranial waveform measurements

**DOI:** 10.1101/2024.01.30.24301974

**Authors:** Faris Gulamali, Pushkala Jayaraman, Ashwin S. Sawant, Jacob Desman, Benjamin Fox, Annie Chang, Brian Y. Soong, Naveen Arivazaghan, Alexandra S. Reynolds, Son Q Duong, Akhil Vaid, Patricia Kovatch, Robert Freeman, Ira S. Hofer, Ankit Sakhuja, Neha S. Dangayach, David S. Reich, Alexander W Charney, Girish N. Nadkarni

## Abstract

**Importance:** Increased intracranial pressure (ICP) is associated with adverse neurological outcomes, but needs invasive monitoring.

**Objective:** Development and validation of an AI approach for detecting increased ICP (aICP) using only non-invasive extracranial physiological waveform data.

**Design:** Retrospective diagnostic study of AI-assisted detection of increased ICP. We developed an AI model using exclusively extracranial waveforms, externally validated it and assessed associations with clinical outcomes.

**Setting:** MIMIC-III Waveform Database (2000-2013), a database derived from patients admitted to an ICU in an academic Boston hospital, was used for development of the aICP model, and to report association with neurologic outcomes. Data from Mount Sinai Hospital (2020-2022) in New York City was used for external validation.

**Participants:** Patients were included if they were older than 18 years, and were monitored with electrocardiograms, arterial blood pressure, respiratory impedance plethysmography and pulse oximetry. Patients who additionally had intracranial pressure monitoring were used for development (N=157) and external validation (N=56). Patients without intracranial monitors were used for association with outcomes (N=1694).

**Exposures:** Extracranial waveforms including electrocardiogram, arterial blood pressure, plethysmography and SpO_2_.

**Main Outcomes and Measures:** Intracranial pressure > 15 mmHg. Measures were Area under receiver operating characteristic curves (AUROCs), sensitivity, specificity, and accuracy at threshold of 0.5. We calculated odds ratios and p-values for phenotype association.

**Results:** The AUROC was 0.91 (95% CI, 0.90-0.91) on testing and 0.80 (95% CI, 0.80-0.80) on external validation. aICP had accuracy, sensitivity, and specificity of 73.8% (95% CI, 72.0%-75.6%), 99.5% (95% CI 99.3%-99.6%), and 76.9% (95% CI, 74.0-79.8%) on external validation. A ten-percentile increment was associated with stroke (OR=2.12; 95% CI, 1.27-3.13), brain malignancy (OR=1.68; 95% CI, 1.09-2.60), subdural hemorrhage (OR=1.66; 95% CI, 1.07-2.57), intracerebral hemorrhage (OR=1.18; 95% CI, 1.07-1.32), and procedures like percutaneous brain biopsy (OR=1.58; 95% CI, 1.15-2.18) and craniotomy (OR = 1.43; 95% CI, 1.12-1.84; *P* < 0.05 for all).

**Conclusions and Relevance:** aICP provides accurate, non-invasive estimation of increased ICP, and is associated with neurological outcomes and neurosurgical procedures in patients without intracranial monitoring.

## Introduction

Elevation in intracranial pressure (ICP) is common in severe acute brain injuries (SABI), such as stroke and traumatic brain injuries, contributing to secondary neurological damage^1–3^. The current gold standard for ICP monitoring is an invasive monitor, which carries risks of infection and hemorrhage, limiting its use^4^.

Non-invasive ICP estimation methods, like transcranial doppler (TCD) and optic nerve sheath diameter (ONSD) show promise for the detection of intracranial hypertension^1,5,6^. However, their utility is constrained by limited availability of specialized skills and equipment settings^7,8^ and the requirement of high clinical suspicion to administer these tests, possibly overlooking subclinical ICP abnormalities. Additionally, there is substantial variability in accuracy and clinical relevance for ICP monitoring using these technologies^9–11^.

Recognizing these limitations, recent research seeks to correlate physiological data with neurological conditions^12^. Yet, such studies are often restricted by their sample size, stringent data filters limiting real world application, and lack of external validation limiting generalizability^12–15^.

To address these limitations, we introduce, a novel method, artificial intelligence derived intracranial pressure (aICP), an approach developed to predict intracranial hypertension. The model utilizes physiologic extracranial waveforms that are routinely collected in intensive care to generate a second-by-second prediction of whether intracranial pressure is elevated^16^. To train the model, we first subset to a patient cohort from a publicly available dataset that had these extracranial waveforms in addition to intraventricular catheters which directly measure intracranial pressure. Second, we use an independent cohort from the Mount Sinai Hospital, to externally validate this approach. Third, we further evaluated the model’s clinical utility in a larger cohort with extracranial waveforms but without invasive intracranial monitoring, examining associations with clinical phenotypes both *a priori* and via unbiased phenome-wide scans. In this cohort, we evaluate clinical implications of undetected intracranial hypertension and aICP by measuring associations with phenotypes that are either a cause or consequence of increased ICP.

## Methods

### Study Setting and datasets

We used single-admission data from two distinct sources: 1) the publicly available MIMIC III Waveform Database Matched Subset^17–19^ contains waveform records from bedside monitors for 10,282 patients admitted to intensive care units at the Beth Israel Deaconess Medical Center (Boston, MA) between 2001 and 2012 (Goldberger et al. 2000; A. E. W. Johnson et al. 2016), and 2) the MSH Bedmaster Matched Database, a database of waveform recordings for 50,894 patients admitted to the Mount Sinai Hospital (New York, NY) between 2018 and 2022 (Figure 1a i). The latter was derived from a deployment of the Bedmaster^TM^ software. This software is engineered to extract and store real-time patient data obtained from networked General Electric Healthcare bedside patient multi-parameter monitors (Excel Medical Electronics, Jupiter, FL).

**Figure 1.**
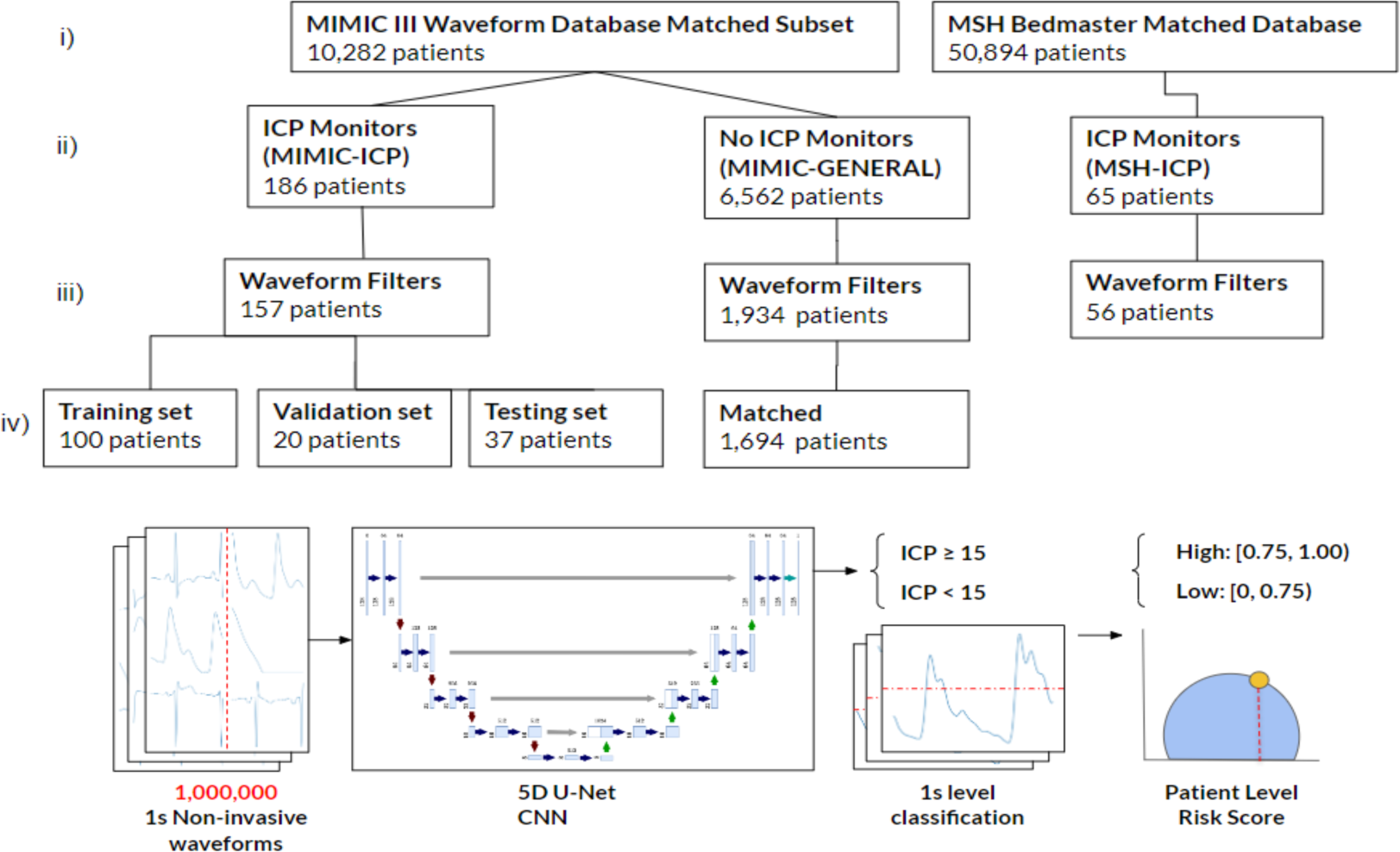
a) Schema for AICP (i) Initial dataset (ii) Layer 1 filters (iii) Layer 2 filters (iv) Final dataset. b) Model Architecture, Training and Output.

We created three cohorts from the two data sources described above: For training and internal validation of aICP, we used part of the MIMIC-III waveform dataset consisting of hospital stays with intracranial waveform recordings (MIMIC-ICP). For external validation of aICP, we used a waveform dataset from Mount Sinai Hospital consisting of hospital stays with intracranial waveform recordings (MSH-ICP). To map aICP to clinical associations, we used another, non-overlapping subset from the MIMIC-III waveform dataset consisting of patients without intracranial waveform recordings (MIMIC-GENERAL) (Figure 1a ii).

### Channels and Data Types Utilized

For all three cohorts, we included arterial blood pressure (ABP), EKG, respiratory, and photoplethysmography (PPG) waveforms. In addition, for MIMIC-ICP and MSH-ICP, we included intracranial pressure waveforms.

### Preprocessing

Since intracranial pressure recordings in both data sources were recorded at 125 Hz, we up-sampled or down-sampled the extracranial waveforms (ABP, EKG, respiratory and PPG) to 125 Hz. We subsequently filtered for quality control (Figure 1a iii). Patient waveforms were included only if they had no missing data and had a non-zero mean and non-zero standard deviation across all waveforms. We found that this procedure removed significant portions of missing or physiologically implausible data that occurs during calibration and setup of the various lines. We randomly subsampled 10,000 1-second segments from each patient.

After filtering, the MIMIC-ICP dataset was partitioned into subsets, with 100 patients allocated to the training dataset, 20 patients to the validation dataset, and 37 patients to the test dataset. The MSH-ICP dataset had 56 patients after filtering (Figure 1a iv). MIMIC-GENERAL had 1,694 after filtering waveforms and EHR data for missingness.

### Thresholding

We employed a threshold of 15 mm Hg to define intracranial hypertension, as previously described in literature^20,21^.

### Model Architecture

aICP utilizes a 5D-convolutional neural network to segment time-series at a second-level^22,23^. These predictions are aggregated to form patient level predictions. Each input consists of the 128 data points (approximately 1 second) of each of the 5 main perioperative waveforms (ABG, EKG leads II and V, respiratory and PPG). The output for each input is a prediction at each second of whether the patient has ICP>15 mm of Hg. Implementation of the model architecture is visualized (**Figure 1B**). The loss function utilized a Tversky Loss, which is a modified DICE score accounting for the relative class imbalance between positive and negative samples^24^.

Briefly, the architecture follows a modified U-Net, a popular neural network design for semantic segmentation tasks, with a modified focus on handling input data with five channels rather than the traditional two or three^25^. It is composed of two convolutional layers with batch normalization and rectified linear unit (ReLU) activation functions, aiming to capture non-linear temporal features within the input data. We add skip connections to handle the high-dimensionality and large number of channels.

Slices within a given time series were labeled using the slice-level prediction model and then transformed into a sequence. The patient-level model aggregated these individual second predictions, generating a unified risk score reflecting the patient’s frequency of dysregulation in intracranial pressure throughout their hospital stay.

### Model Development and Validation

The aICP model has approximately 10.4 million parameters and was trained on a single NVIDIA A100 GPU over the span of 3 days and 30 epochs. The segmentation performance was optimized using a modified Dice coefficient. The optimal model was selected and calibrated based on its performance on the validation dataset. Additionally, we employed hyperparameter optimization, early stopping and an Adam optimizer within the PyTorch Lightning framework.

### Model Performance Metrics

We assessed the classification performance of time series and patient-level data using five key metrics: three threshold-dependent metrics: accuracy, sensitivity, specificity, and two threshold-independent metrics: area-under-the-receiver-operator-curve (AUROC) and area-under-the-precision-recall curve (AUPRC). Receiver operating curves (ROCs) and confusion matrices evaluated the patient-level model performance in each classification task. The threshold was calibrated on the internal validation set, and then applied to the internal test set and external validation set.

To assess the performance of our model, we conducted a benchmark comparison against standard time series classification models. They included Recurrent Neural Networks (RNNs), Long Short-Term Memory networks (LSTMs), and Temporal Convolutional Neural Networks (TCNs), which are state-of-the-art (Nair et al. 2023). All experiments were seeded, and our models underwent consistent training, validation, and testing on identical datasets bootstrapped 50 times (**Supplemental Table 2**).

### Association with Phenotypes and Outcomes

To evaluate the association of aICP with relevant clinical outcomes in patients without intracranial pressure monitors, we used a testing set consisting of patients who had only extracranial waveforms (ABP, EKG, respiratory, PPG) in addition to clinical data from the electronic health record. aICP provided a patient level risk score for 1,694 patients. We calculated an odds ratio with respect to a ten percentile increase increase in aICP. We ran a phenome-wide association and reported the P-values and as well as the adjusted threshold for correlated phenotypes (P < 3.33 × 10^−4^). We then stratified patients into low-and high-risk groups using the 75th percentile of predicted patient level risk score as the cutoff, and calculated odds ratios for pre-determined outcomes. P-value was calculated using the Fisher’s Exact Test.

### Statistical Analysis

We calculated descriptive statistics for each cohort by mapping waveform IDs to electronic health records for available patients. To quantify the uncertainty in experimental results, confidence intervals were computed using via non-parametric bootstrapping 50 times. All evaluation and statistical metrics were computed utilizing the torchmetrics and statsmodels packages^26,27^. Odds ratios were calculated by logistic regression and Fisher’s Exact Test. We additionally fill out the TRIPOD reporting guideline and include it in the Supplement (**Supplementary Figure 1**).

## Results

### Cohort Description

Average ages were 67.7 for the MIMIC-ICP. Females accounted for 40.8% of the MIMIC-ICP cohort, and 60.5% identified as White. In the MIMIC-ICP cohort, 53.3% of patients were on Medicare, 23.3% on private insurance, and 20.0% on Medicaid. The MIMIC-ICP cohort exhibited an average intracranial blood pressure of 8.9 mmHg, and intracranial hypertension values were observed for 9.7% of total observation period in the MIMIC-ICP cohort. Neurological comorbidities were documented for all patients in both MIMIC-ICP, but only 19% of the MIMIC-ICP had documented cardiovascular comorbidities as defined by phecodes^28^. Cardiovascular comorbidities in our study encompassed conditions such as ischemic heart disease, valvular disease, and arrhythmias. Additionally, neurological comorbidities were classified into categories including vascular etiologies (e.g., intracerebral hemorrhage), infectious etiologies (e.g., abscesses), and oncological issues (e.g., tumors) (**Supplementary Table 3**).

56 patients admitted to the Mount Sinai Hospital between 2020 and 2022 with intracranial monitors comprised MSH-ICP. The average age was 54.6 (SD, 31.3) with 39.3% of patients identifying as female. 50.0% of patients identified as White, 16.1% as Black or African American, 17.8% as Hispanic, with the remaining 14.3% identifying as other. 28.6% of patients were on Medicare, 42.8% on private insurance and 28.6% on Medicaid. This cohort demonstrated an average intracranial pressure of 16.7 mmHg. Elevated ICP was observed for a total of 8% of the total observation period. All patients in this cohort had neurological and cardiovascular comorbidities.

For the clinical association cohort (MIMIC-GENERAL), the average age was 69.3 years and 41% of the patients were female. 71.0% of patients identified as White, 10.0% as Black or African American, 2.7% as Hispanic and 16.3% as other. 55.4% were on Medicare, 30.3% on private insurance and 10.7% on Medicaid. 52.8% of patients had an identifiable neurological comorbidity and 56.1% had an identified cardiovascular comorbidity.

### aICP Performance

First, we assessed the patient-level performance of aICP using the testing set (**Table 2**). The Receiver Operating Characteristic (ROC) curve was employed to measure the model’s ability to detect elevated intracranial pressure (ICP > 15 mm Hg), with overall accuracy of 0.97, and an area-under-the-curve (AUC) of 0.91. The sensitivity of the model at a threshold of 0.5 was 1.00 and the specificity was 0.87. These results outperformed existing models (Supplementary Table 2).

**Table 1.** Patient Characteristics.

|  | MIMIC-ICP<br>With ICP<br>monitors<br>(n=157) | MIMIC-GENERAL<br>Without ICP<br>monitors<br>(n=1694) | MSH-ICP<br>With ICP<br>monitors<br>(n=56) | P |
| --- | --- | --- | --- | --- |
| Mean Age, (SD) | 67.7<br>(13.3) | 69.3 (13.8) | 54.6<br>(31.3) | <0.01 |
| Female, % | 40.8 | 41 | 39.3 |  |
| Self-Reported Race/Ethnicity, % |  |  |  |  |
| White | 60.5 | 71.0 | 50.0 | < 0.01 |
| Black / African American | 3 | 10 | 16.1 |  |
| Non-White Hispanic | 3 | 2.7 | 17.8 |  |
| Other | 34.5 | 16.3 | 14.3 |  |
| Insurance, % |  |  |  |  |
| Medicare | 53.3 | 55.4 | 28.6 | <0.01 |
| Private | 23.3 | 30.3 | 42.9 |  |
| Medicaid | 20.0 | 10.7 | 28.6 |  |
| Other | 3.4 | 3.6 | 0 |  |
| Measured intracranial Pressure in mm of Hg, Mean (SD) | 8.9 (7.3) | NA | 16.7 (5.2) | <0.01 |
| Measured intracranial hypertension, % | 9.7 | NA | 8.0 | <0.01 |
| Neurological Disease, % | 100 | 52.8 | 100 | <0.01 |
| Cardiovascular Disease, % | 19 | 56.1 | 100 | <0.01 |

P values for continuous values are calculated via ANOVA, and for binary variables are calculated via Chi-Squared test. Neurological and Cardiovascular outcomes are defined by ICD-9 codes (**Supplementary Table 3**).

**Table 2.** Performance of patient level classification model on internal and external testing set, evaluated with accuracy thresholded at 0.5, sensitivity thresholded at 0.5, specificity thresholded at 0.5, area under the receiving-operator-curve, and area under the precision-recall curve for ICP elevation.

|  | MIMIC-ICP | MSH-ICP |
| --- | --- | --- |
| Accuracy (95% CI) | 0.972 (0.971-0.973) | 0.738 (0.720-0.756) |
| Area Under the Receiver Operating Curve (95% CI) | 0.91 (0.90-91) | 0.80 (0.79-0.80) |
| Area Under the Precision Recall Curve (95% CI) | 0.91 (0.91-0.91) | 0.93 (0.93-0.93) |
| Sensitivity (95% CI) | 1.0 (1.0-1.0) | 0.99 (0.99-0.99) |
| Specificity (95% CI) | 0.87 (0.86-0.87) | 0.77 (0.74-0.79) |

Next, we evaluated the performance on MSH-ICP, external validation test set. The area-under-the-curve on the external validation test set was 0.80, with an accuracy of 0.74, a sensitivity of 0.99 and a specificity of 0.77. The Receiver Operating Curve and Precision-Recall Curves for internal and external validation testing sets showed discriminatory performance of aICP to identify patients with ICP values greater than 15mmHg (**Figure 2**).

**Figure 2.**
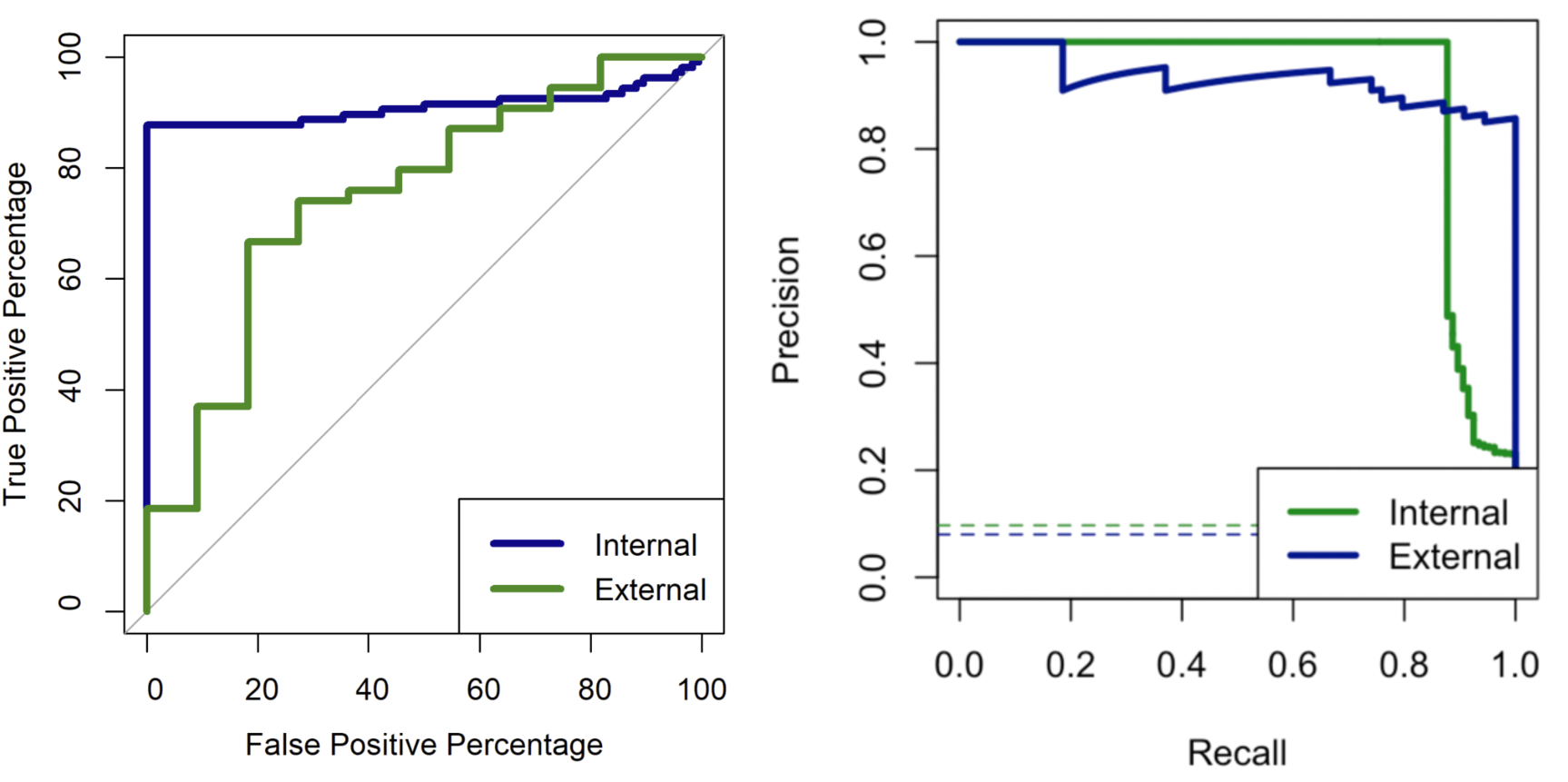
a) AUROC curve on the internal (MIMIC-ICP) and external (MSH-ICP) test cohorts. b) AUPRC curve for internal (MIMIC-ICP) and external (MSH-ICP) test cohorts.

To evaluate the association of aICP with relevant clinical outcomes in patients without intracranial pressure monitors, we used MIMIC-GENERAL, a testing set consisting of patients that had extracranial waveforms (ABP, EKG, respiratory and PPG) and linked clinical data. aICP provided a patient level risk score for 1,694 patients. We calculated an odds ratio with respect to a 10 percentile increase increase in aICP (Figure 3A). A ten-percentile increase in aICP was associated with an increased likelihood of stroke (OR=2.12; 95% CI, 1.27-3.13; *P*=4.06×10^−3^), brain malignancy (OR=1.68; 95% CI, 1.09-2.60; *P*=1.93×10^−2^), subdural hemorrhage (OR=1.66; 95% CI, 1.07-2.57; P=2.38×10^−2^), intracerebral hemorrhage (OR=1.18; 95% CI, 1.07-1.32; *P*=1.17×10^−3^). Moreover, when looking at patient procedures done over the course of admission, a ten percentile increase in aICP was associated with a percutaneous brain biopsy (OR=1.58; 95% CI, 1.15-2.18; *P*=4.58×10^−3^), and craniotomy and resection (OR=1.43; 95% CI, 1.12-1.84; *P*=4.10×10^−3^) (Figure 3A).

**Figure 3.**
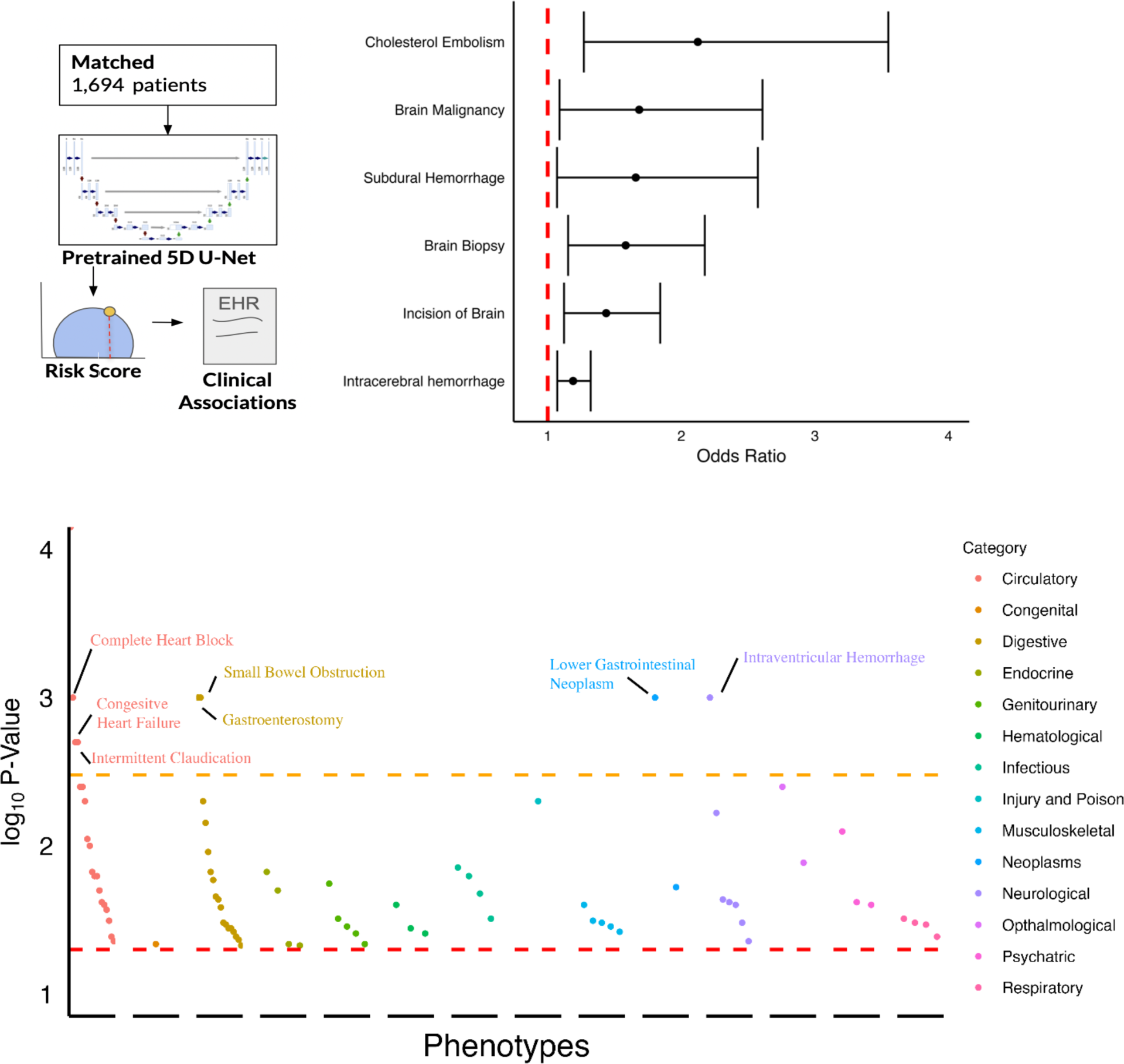
a) Schema depicting the phenotype wide association scan between aICP and phenotypes. b) A decile increase in aICP is associated with various increases in odds ratios of having been previously or currently being diagnosed with a neurovascular condition. c) Unadjusted P-values from a phenome wide scan of aICP across different categories colored and sized by odds ratio with the red dashed line representing the unadjusted threshold and the orange dashed line representing the adjusted threshold.

To see if the effect of aICP was larger in high-risk groups, we then stratified patients into low and high-risk groups, using the 75^th^ percentile of predicted patient level risk score as the cutoff. We calculated odds ratios and P-values via the Fisher’s Exact Test for pre-determined outcomes. Patients in the top quartile demonstrated increased risk of subdural hemorrhage (OR=24.2; *P=3.01×10^−2^*), traumatic brain injury (OR=6.04; *P=3.8 ×10^−2^*), intracerebral hemorrhage (OR=1.85; *P=1.32×10^−3^*), and receiving a craniectomy (OR=7.55; *P=1.58×10^−2^*) or a percutaneous brain biopsy (OR=5.03; *P=2.72×10^−2^*).

### Phenome-wide association discovery

Finally, we sought to discover whether elevated aICP was correlated with other pathologies. We mapped ICD-9 codes to phenotypic groups via PheCodes^28^. We ran a phenotype-wide association scan and demonstrated that belonging to the top quartile of an aICP-derived risk score was strongly associated with circulatory pathologies (N=13), acute liver failure (N=9), metabolic abnormalities and chronic kidney disease (N=6). For example, an elevated aICP was linked to myocardial infarction (OR=1.75; *P*=2.65×10^−3^), valvular disease (OR=2.07; *P*=2.88×10^−2^), and peripheral vascular disease with claudication (OR=4.24; *P*=1.39×10^−2^). In the context of liver failure, an aICP-related risk stratification demonstrated associations with ascites (OR=1.93; *P=*5.68×10^−3^), esophageal varices (OR=2.99; *P*=1.22×10^−3^), and portal vein thrombosis (OR=4.05; *P*=4.00×10^−2^). In the context of metabolic abnormalities and acute renal failure, aICP is associated with diabetes mellitus without complications (OR=1.34; *P*=1.62×10^−2^), diabetes mellitus with renal involvement (OR=2.08; *P*=2.88×10^−2^), and with malignant hypertension from kidney failure with uncontrolled diabetes (OR=18.2; *P*=1.32×10^−3^). Finally, other notable associations with elevated aICP include glaucoma (OR=2.72; *P*=4.68×10^−3^) and changes in vision (OR=1.73; *P*=1.32×10^−3^) (**Supplementary Table 4)**.

## Discussion

We introduce a novel deep learning system named aICP, that serves as a digital biomarker of intracranial hypertension. We developed and validated aICP both internally and externally for ICP greater than 15 mmHg in patients with invasive ICP monitors. In patients without ICP monitoring, we show that aICP is strongly associated with relevant neurological phenotypes, indicating that it may be able to serve as a screening tool in patients not subjected to intracranial monitoring but who have extracranial monitors.

aICP demonstrated an AUROC of 80% in MSH-ICP, the external validation cohort. It demonstrated promise as a screening tool, with high sensitivity (99.5%) in the external validation dataset (99.5%) at a threshold of 0.5. The slight drop in performance compared to internal validation cohort may be related to the demographic differences in the cohorts, as well as differences in monitoring devices and neurosurgical practices. The training and validation cohorts exhibited comparable distributions of sex and prevalence of neurological disorders. However, there were significant differences in racial, ethnic and insurance distributions across the cohorts. Additionally, patients in MSH-ICP demonstrated more disease severity, indicated by a significantly higher average intracranial blood pressure compared to patients in MIMIC-ICP. Moreover, all patients in the MSH-ICP had documented circulatory disorders, in contrast to the MIMIC-ICP cohort, where only 20% of patients had recorded cardiovascular phenotypes. Despite these differences, which may influence EKG and arterial line signals, the performance remained robust in the external validation cohort.

In patients who did not have ICP monitors, aICP demonstrated associations with specific neurologic pathologies and neurosurgical procedures. These associations suggest aICP’s clinical value in identifying high-risk patients who may benefit from enhanced monitoring. Notably, patients in the top quartile exhibited significantly elevated risks for subdural hemorrhage, traumatic brain injury, intracerebral hemorrhage, craniectomy, and percutaneous brain biopsy, indicating the tool’s potential to guide tailored interventions.

Thus, we provide two specific potential clinical use cases for aICP. First, aICP can provide intracranial monitoring to patients that have contraindications to invasive monitoring. For example, external ventricular drain placement is contraindicated in patients on anticoagulation due to bleeding risk. In patients on anticoagulation, aICP can be used in place of an external ventricular drain to monitor for ischemic-to-hemorrhagic conversion of a stroke. Second, because aICP needs only data from routine extracranial monitors, it could be utilized as early as arrival to the emergency department in the context of traumatic brain injury.

A comprehensive exploration across non-neurological phenotypes revealed associations of aICP with renal failure and acute liver failure, which may provide insight into the etiology of hepatic and uremic encephalopathy and consequently increased ICP^29,30^. Additionally, aICP demonstrated associations with glaucoma, as evidenced by significant correlations between elevated intracranial and intraocular pressures^31^. These findings highlight the multifaceted potential of aICP in providing clinical insights through diverse associations beyond neurovascular domains, making it an asset for comprehensive patient assessment.

As with all clinical tools, our method also has limitations. This is, to our knowledge, one of the largest clinical datasets used to train a deep learning algorithm for intracranial pressure estimation. While our clinical validation cohort (MSH-ICP) is very diverse in race and ethnicity, as it is reflective of the current diversity of New York City, it has fewer patients than the MIMIC-ICP (training) cohort. The significant difference in sample distribution between MIMIC-III and MSH-ICP may affect the interpretation of our results. However, awareness of disparities in racial, ethnic, and socioeconomic backgrounds is crucial for understanding how results may generalize to different patient populations. Some patients had to be excluded when we encountered significant patient drift in the measured variables. Prospective studies with large, diverse patient cohorts will be necessary to explore the practical implementation of aICP.

In summary, we developed aICP to detect intracranial pressure abnormalities demonstrating both strong performance and clinical significance. aICP does not have any non-standard hardware requirements, and consequently, implementation of aICP as a tool for point-of-care bedside monitoring at the start of admission can occur on routine bedside monitors. Moreover, an aICP-generated risk score for each patient over the course of a patient’s hospital stay, could reduce detection times, and improve outcomes, especially given the time-sensitive nature of neurological diseases such as stroke and hemorrhage. Finally, we anticipate that aICP has the potential to detect pathophysiological conditions like hepatic encephalopathy and glaucoma, which are associated with intracranial pressure changes but do not have clear neurovascular etiologies.

## Data Availability

Data from the publicly available MIMIC-III dataset can be obtained by registering at physionet.org. Data from Mount Sinai Hospital contains private health information and we do not make this available due to privacy concerns.

## Data Sharing

Data can be obtained via the MIMIC-III online repository. MSH-ICP data is from the Mount Sinai Data Warehouse but contains private health information. We do not make this data publicly available due to concerns for healthcare data privacy.

## Acknowledgements

This work was supported in part through the computational and data resources and staff expertise provided by Scientific Computing and Data at the Icahn School of Medicine at Mount Sinai and supported by the Clinical and Translational Science Awards (CTSA) grant UL1TR004419 from the National Center for Advancing Translational Sciences. Research reported in this publication was also supported by the Office of Research Infrastructure of the National Institutes of Health under award number S10OD026880 and S10OD030463. The content is solely the responsibility of the authors and does not necessarily represent the official views of the National Institutes of Health.

## Supplementary Materials

**Supplementary Table 1.** Comparison of Various Methods Supplementary Table 1.

| Model | Sample Size | Data | Reported AUROC | External | Clinical |
| --- | --- | --- | --- | --- | --- |
| RF <sup>4</sup> | 110 | BOOST-2 | 0.84 | No | Yes |
| B4C <sup>5</sup> | 40 | Sao Paulo | 0.90 | No | No |
| TCD <sup>6</sup> | 262 | IMPRESSIT-2 | 0.76 | No | No |
| TCD <sup>7</sup> | 11 | Columbia | N/A | No | No |
| EKG+ART <sup>8</sup> | 10 | MIMIC | N/A | No | No |
| ONSD <sup>9</sup> | 100 | University of Libre Bruxelles | 0.85 (0.77-0.93) | No | No |
| JVP <sup>10</sup> | 11 | VGH | 0.75 (0.56-0.94) | No | No |
| ONSD+JVP <sup>10</sup> | 11 | VGH | 0.91 (0.84-0.97) | No | No |
| **aICP** | 212 | MIMIC | 0.91 (0.90, 0.92) | Yes | 1,694 |

**Supplementary Table 2.**
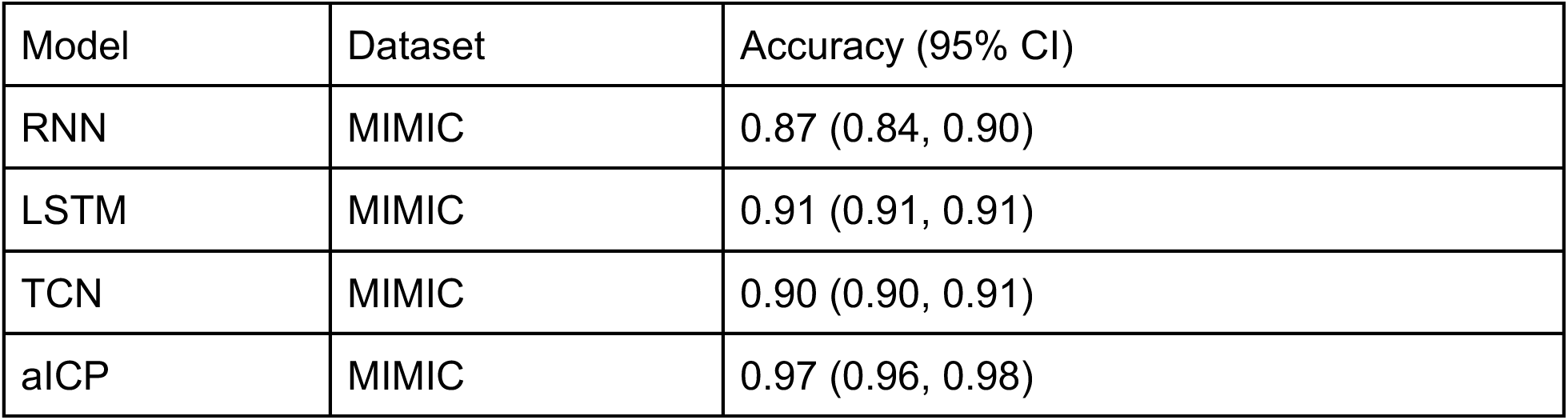
Benchmarking against other ML methods.

| Model | Dataset | Accuracy (95% CI) |
| --- | --- | --- |
| RNN | MIMIC | 0.87 (0.84, 0.90) |
| LSTM | MIMIC | 0.91 (0.91, 0.91) |
| TCN | MIMIC | 0.90 (0.90, 0.91) |
| aICP | MIMIC | 0.97 (0.96, 0.98) |

**Supplementary Table 3.**
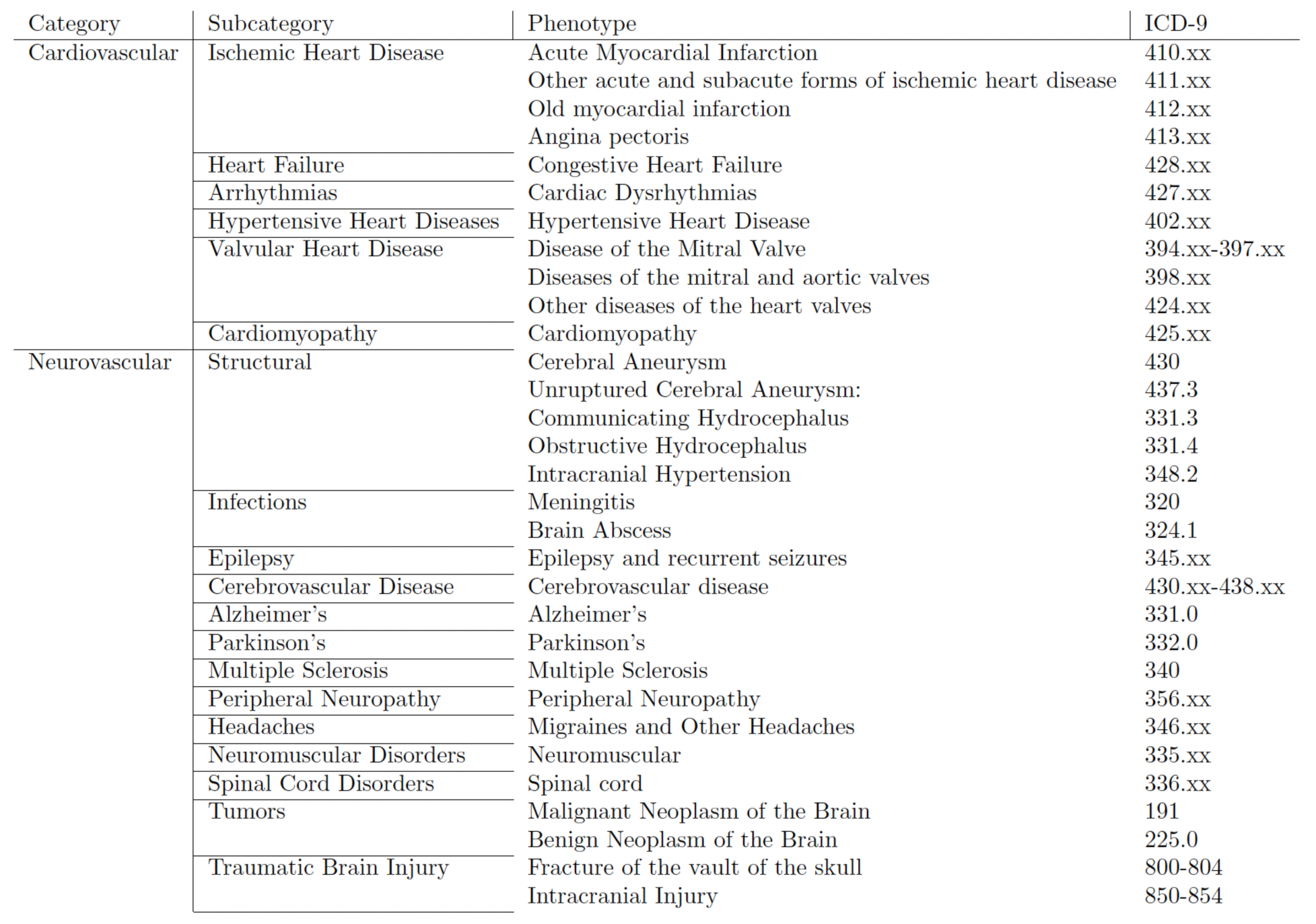
Categorization of Phenotypes for Demographic Analysis.

**Supplementary Table 4.** Phenome Wide Scan.

| Phenotype | P-value | odds ratio | Lower CI | Upper CI | Group |
| --- | --- | --- | --- | --- | --- |
| Atrioventricular block, complete | 0.000 | 1.35 | 1.14 | 1.59 | Circulatory |
| Malignant neoplasm of rectum, rectosigmoid junction, and anus | 0.001 | 1.73 | 1.27 | 2.36 | Neoplasms |
| Congestive Heart Failure | 0.001 | 1.12 | 1.05 | 1.20 | Circulatory |
| Small Bowel Obstruction | 0.001 | 1.44 | 1.16 | 1.79 | Digestive |
| Gastroenterostomy | 0.001 | 1.27 | 1.10 | 1.46 | Digestive |
| Intracerebral hemorrhage | 0.001 | 1.19 | 1.07 | 1.32 | Neurological |
| Atherosclerosis of native arteries of the extremities with intermittent claudication | 0.00<br>2 | 1.53 | 1.17 | 2.00 | Circulatory |
| Subendocardial Infarction | 0.00<br>2 | 1.17 | 1.06 | 1.30 | Circulatory |
| Glaucoma | 0.00<br>4 | 1.31 | 1.09 | 1.57 | Ophthalmological |
| Cholesterol Embolism | 0.00<br>4 | 2.12 | 1.27 | 3.55 | Circulatory |
| Pacemaker | 0.00<br>4 | 1.14 | 1.04 | 1.25 | Circulatory |
| Incision uterine septum | 0.00<br>5 | 0.40 | 0.21 | 0.76 | Circulatory |
| Ascites (non malignant) | 0.00<br>5 | 0.80 | 0.68 | 0.93 | Digestive |
| Aromatic Analgesic Poisoning | 0.00<br>5 | 0.00 | 0.00 | 0.01 | Injury and Poison |
| Epilepsy, recurrent seizures, convulsions | 0.00<br>6 | 1.28 | 1.07 | 1.53 | Neurological |
| Partial esophagectomy | 0.00<br>7 | 1.19 | 1.05 | 1.35 | Digestive |
| Anxiety | 0.00<br>8 | 0.76 | 0.61 | 0.93 | Psychiatric |
| Atrial fibrillation | 0.00<br>9 | 1.09 | 1.02 | 1.17 | Circulatory |
| CABG | 0.01<br>0 | 1.30 | 1.07 | 1.58 | Circulatory |
| Truncal vagotomy | 0.01<br>1 | 1.47 | 1.09 | 1.98 | Digestive |
| Disturbances of vision | 0.01<br>3 | 1.73 | 1.12 | 2.67 | Ophthalmological |
| Postoperative infection | 0.01<br>4 | 0.76 | 0.61 | 0.95 | Infectious |
| Heart Failure | 0.01<br>5 | 1.40 | 1.07 | 1.84 | Circulatory |
| Diabetes without Complications | 0.01<br>5 | 1.09 | 1.02 | 1.17 | Endocrine |
| Other tests | 0.01<br>5 | 0.00 | 0.00 | 0.04 | Digestive |
| Anticoagulants causing adverse effects | 0.01<br>6 | 1.22 | 1.04 | 1.43 | Circulatory |
| Methicillin resistant Staphylococcus aureus | 0.01<br>6 | 1.45 | 1.07 | 1.96 | Infectious |
| Atherosclerosis of the extremities | 0.01<br>6 | 1.32 | 1.05 | 1.65 | Circulatory |
| Perforated Intestine | 0.01<br>7 | 0.36 | 0.16 | 0.83 | Digestive |
| Bladder Obstruction | 0.01<br>8 | 0.80 | 0.67 | 0.96 | Genitourinary |
| Secondary malignancy of brain/spine | 0.01<br>9 | 1.68 | 1.09 | 2.61 | Neoplasms |
| Hypopotassemia | 0.02<br>0 | 0.80 | 0.66 | 0.97 | Endocrine |
| Hemorrhage or hematoma complicating a procedure | 0.02<br>0 | 0.81 | 0.68 | 0.97 | Circulatory |
| Long-Term Antibiotics | 0.02<br>1 | 1.67 | 1.08 | 2.59 | Infectious |
| Peptic Ulcer | 0.02<br>2 | 1.59 | 1.07 | 2.38 | Digestive |
| Proctosigmoidoscopy | 0.02<br>3 | 1.33 | 1.04 | 1.69 | Digestive |
| Dental scaling & debride | 0.02<br>3 | 0.01 | 0.00 | 0.53 | Neurological |
| Cannabis Use | 0.02<br>4 | 0.00 | 0.00 | 0.15 | Psychiatric |
| Subdural Hemorrhage | 0.02<br>4 | 1.66 | 1.07 | 2.57 | Neurological |
| Disease of tricuspid valve | 0.02<br>4 | 1.24 | 1.03 | 1.49 | Circulatory |
| Calcaneal spur; Exostosis NOS | 0.02<br>5 | 0.00 | 0.00 | 0.17 | Musculoskeletal |
| Vascular Grat | 0.02<br>5 | 1.28 | 1.03 | 1.59 | Circulatory |
| Alcohol Use Disorder | 0.02<br>5 | 0.67 | 0.47 | 0.95 | Psychiatric |
| Vocal Cord Paralysis | 0.02<br>5 | 1.58 | 1.06 | 2.36 | Neurological |
| Essential thrombocythemia | 0.02<br>5 | 0.57 | 0.35 | 0.93 | Hematological |
| Fluid overload | 0.02<br>6 | 0.76 | 0.60 | 0.97 | Digestive |
| Systolic Heart Failure | 0.02<br>7 | 1.17 | 1.02 | 1.34 | Circulatory |
| Vesicostomy | 0.03<br>1 | 0.78 | 0.62 | 0.98 | Genitourinary |
| Shortness of breath | 0.03<br>1 | 0.00 | 0.00 | 0.45 | Respiratory |
| Fever of unknown origin | 0.03<br>1 | 1.25 | 1.02 | 1.54 | Infectious |
| Acute Congestive Heart Failure<br>Exacerbation | 0.03<br>2 | 1.15 | 1.01 | 1.32 | Circulatory |
| Osteoclasia | 0.03<br>2 | 1.95 | 1.06 | 3.59 | Musculoskeletal |
| Disorders of muscle, ligament, and<br>fascia | 0.03<br>3 | 1.42 | 1.03 | 1.96 | Musculoskeletal |
| Ptosis | 0.03<br>3 | 1.62 | 1.04 | 2.52 | Neurological |
| Lung Collapse | 0.03<br>3 | 1.55 | 1.04 | 2.33 | Respiratory |
| Diverticulitis | 0.03<br>3 | 0.46 | 0.23 | 0.94 | Digestive |
| Painful respiration | 0.03<br>4 | 1.94 | 1.05 | 3.56 | Respiratory |
| Esophageal bleeding<br>(varices/hemorrhage) | 0.03<br>4 | 0.73 | 0.55 | 0.98 | Digestive |
| Urinary incontinence | 0.03<br>5 | 1.93 | 1.05 | 3.54 | Genitourinary |
| Fracture of ribs | 0.03<br>5 | 1.71 | 1.04 | 2.83 | Musculoskeletal |
| Colostomy | 0.03<br>6 | 1.36 | 1.02 | 1.82 | Digestive |
| Deep Vein Thrombosis | 0.03<br>6 | 1.39 | 1.02 | 1.90 | Hematological |
| Vagotomy | 0.03<br>6 | 1.39 | 1.02 | 1.90 | Digestive |
| Hematemesis | 0.03<br>8 | 0.46 | 0.22 | 0.96 | Digestive |
| Intertrochanteric Fracture | 0.03<br>8 | 1.41 | 1.02 | 1.94 | Musculoskeletal |
| Urethral Stricture | 0.03<br>9 | 1.70 | 1.03 | 2.81 | Genitourinary |
| Phlebitis and thrombophlebitis | 0.03<br>9 | 1.53 | 1.02 | 2.31 | Hematological |
| Inferoposterior Acute Myocardial<br>Infarction | 0.04<br>1 | 0.00 | 0.00 | 0.63 | Circulatory |
| Thoracic Esophagocenteresis | 0.04<br>1 | 1.14 | 1.01 | 1.28 | Digestive |
| Tracheostomy | 0.04<br>1 | 0.87 | 0.76 | 0.99 | Respiratory |
| Peritoneal Injury | 0.04<br>3 | 1.87 | 1.02 | 3.44 | Digestive |
| Cardiac Catheterization | 0.04<br>4 | 1.18 | 1.00 | 1.40 | Circulatory |
| Sphenoidotomy | 0.04<br>4 | 0.72 | 0.53 | 0.99 | Neurological |
| Urinary Retention | 0.04<br>6 | 1.17 | 1.00 | 1.37 | Genitourinary |
| Down Syndrome | 0.04<br>6 | 1.52 | 1.01 | 2.29 | Congenital |
| Hypertensive chronic kidney disease | 0.04<br>6 | 1.44 | 1.01 | 2.06 | Endocrine |
| Peritonitis and retroperitoneal infections | 0.04<br>7 | 0.69 | 0.48 | 0.99 | Digestive |
| Dysthymic disorder | 0.04<br>7 | 0.79 | 0.63 | 1.00 | Endocrine |

**Supplementary Figure 1.**
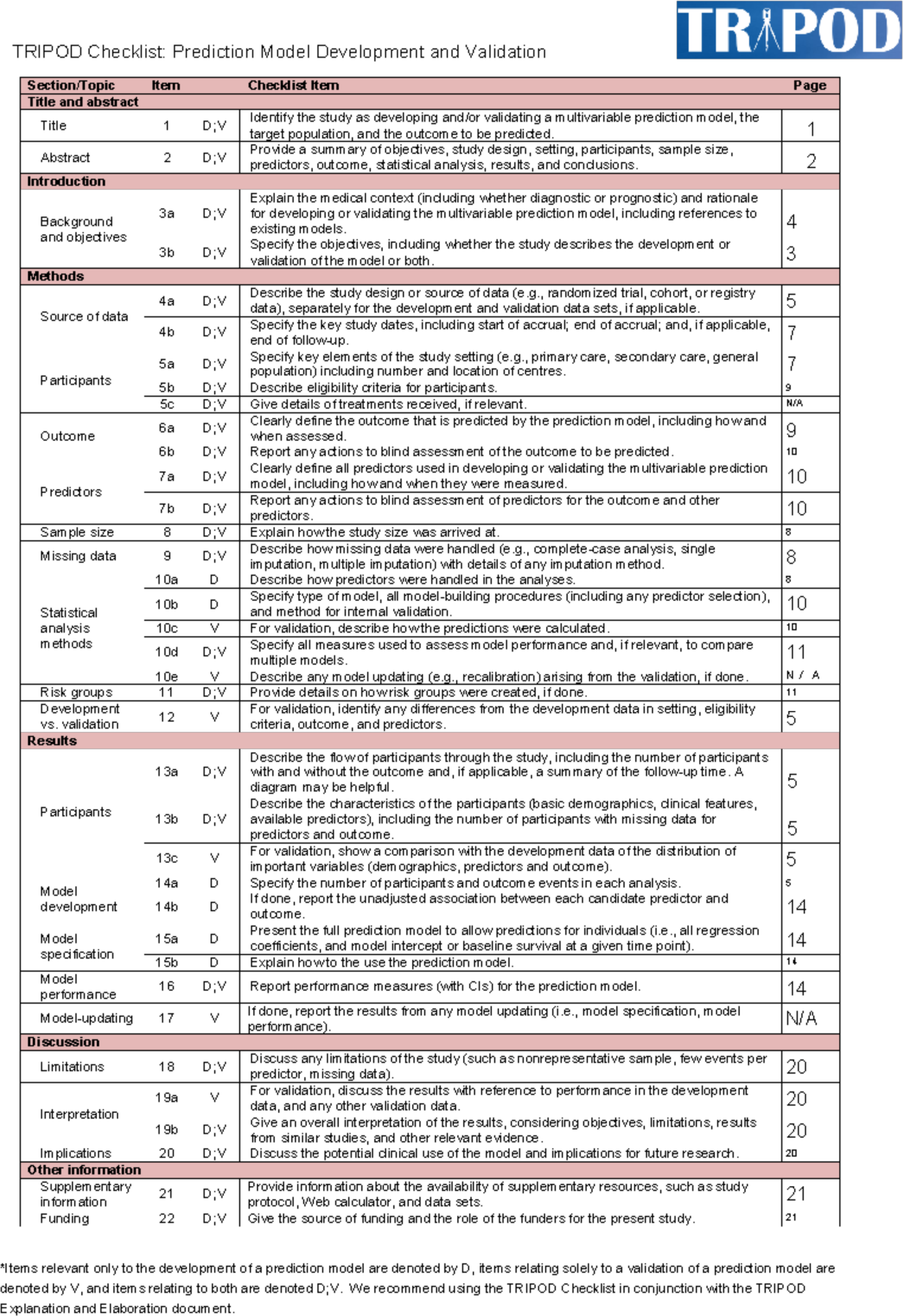
TRIPOD Checklist.

